# An epithelial microRNA upregulates airway IL-25 and TSLP expression in type 2-high asthma via targeting CD39-extracellular ATP axis

**DOI:** 10.1101/2020.09.07.20188987

**Authors:** Kan Zhang, Yuchen Feng, Yuxia Liang, Wenliang Wu, Chenli Chang, Dian Chen, Shengchong Chen, Jiali Gao, Gongqi Chen, Lingling Yi, Dan Cheng, Guohua Zhen

## Abstract

**Introduction:** Epithelial cell-derived cytokines IL-25, IL-33 and TSLP initiate type 2 inflammation in allergic diseases. However, the signaling pathway regulating these cytokines expression remains elusive. We examined the role of epithelial microRNAs in the expression of IL-25, IL-33 and TSLP in asthma.

**Methods:** Differentially expressed epithelial microRNAs between type 2-low and type 2-high asthma patients were identified using microarray. The expression of microRNA (miR)-206, its target CD39, CD39’s substrate ATP, and IL-25, IL-33, TSLP were measured in epithelial brushings and bronchoalveolar lavage fluid from both asthma subsets and healthy controls. The links between these measurements were functionally validated in vitro and in vivo.

**Results:** MiR-206 was the most highly expressed microRNA in type 2-high asthma relative to type 2-low asthma, but was downregulated in both asthma subsets compared with healthy controls. CD39, an ecto-nucleotidase degrading ATP, was a target of miR-206 and upregulated in asthma. Allergen-induced acute extracellular ATP accumulation led to miR-206 downregulation and CD39 upregulation in human bronchial epithelial cells, forming a feedback loop to eliminate excessive ATP. Airway ATP levels strongly correlated with elevated IL-25 and TSLP expression in type 2-high asthma patients. Intriguingly, airway miR-206 antagonism increased Cd39 expression, reduced ATP accumulation, suppressed Il-25, Il-33, Tslp expression and group 2 innate lymphoid cells expansion, and alleviated type 2 inflammation in a mouse model of asthma. However, airway miR-206 overexpression had opposite effects.

**Conclusion:** Together, epithelial miR-206 upregulates airway IL-25, TSLP expression via targeting CD39-extracellular ATP axis, which represents a novel therapeutic target in type 2-high asthma.

## INTRODUCTION

Asthma is a heterogeneous disease with different phenotypes, endotypes, and variable responses to management approaches [1, 2]. A prominent endotype of asthma is the presence of type 2 inflammation [3, 4]. Airway epithelial cells play pivotal roles in the initiation of type 2 inflammation by expressing interleukin (IL)-25, IL-33 and thymic stromal lymphopoietin (TSLP) [5-7]. These epithelial cell-derived cytokines act on innate immune cells including dendritic cells, group 2 innate lymphoid cells (ILC2s) and mast cells [8-15]. Recent identification of memory Th2 cells with high expression of receptor for IL-25, IL-33 and TSLP supports a role of these cytokines in adaptive immune responses in allergy [16, 17]. IL-25 [18, 19], IL-33 [20], and TSLP [11] have each been reported to be indispensable and sufficient for type 2 cytokine production, eosinophilic airway inflammation and AHR in certain mouse models. In human asthma, various expression patterns of these cytokines have been reported [21-25]. Although IL-25, IL-33 and TSLP are critical in type 2 airway inflammation, the upstream signaling pathway regulating their expression remains elusive.

When exposed to environmental stimuli, airway epithelial cells rapidly express danger signals such as adenosine triphosphate (ATP), uric acid to alert the immune system [26, 27]. Allergen exposure enhanced airway ATP concentration in human asthma and sensitized mice. Neutralizing airway ATP or blocking of purine signaling suppressed airway inflammation in allergen sensitized and challenged mice [28]. Extracellular ATP concentration is tightly controlled by ecto-nucleoside triphosphate diphosphohydrolases (ENTPD) [29]. CD39 (encoded by *ENTPD1*), catalyzing the degradation of extracellular ATP and ADP, is expressed in airway epithelial cells [30]. Inhibition of CD39 activity increased BALF ATP concentration and intensified ovalbumin-induced bronchospasm in mice [31]. CD39-deficient mice exhibited worsened airway inflammation and mucus overproduction after allergen sensitization and challenge [32], whereas the paradoxically alleviated airway inflammation was also reported in CD39-dificient mice [33]. Recently, it is reported that ATP serves as a sensor for an airborne allergen to trigger IL-33 release in airway epithelial cells [34]. Together, these studies suggest that CD39-extracellular ATP axis may regulate IL-25, IL-33 and TSLP in asthma.

MicroRNAs (miRNAs) regulate gene expression by promoting the catabolism of the target transcripts as well as attenuating their translation. There are a growing body of evidence that miRNAs play important roles in epithelial cell, ILC2 and Th2 cell differentiation and function in asthma [35-38]. MiR-19a promotes IL-5, IL-13 expression in ILC2s and Th2 cells [37, 38]. Let-7 miRNA regulates IL-13 expression and allergic airway inflammation [39, 40]. The global miRNAs expression in airway epithelial cells are altered in asthma [36, 41]. However, the difference of epithelial miRNA expression between type 2-low and type 2-high asthma remains unknown. We hypothesized that the differentially expressed epithelial miRNAs between the two asthma subsets may contribute to IL-25, IL-33 and TSLP expression in asthma.

In this study, we profiled the epithelial miRNAs’ expression in type 2-low and -high asthma patients. We found that miR-206, the most highly expressed miRNA in type 2-high asthma relative to type 2-low asthma, targets CD39-extracelluar ATP axis to regulate IL-25 and TSLP expression in cultured bronchial epithelial cells. Airway ATP levels were increased and strongly correlated with elevated IL-25 and TSLP expression in type 2-high asthma patients. In a mouse model of asthma, manipulation of airway miR-206 expression altered Il-25, Il-33, Tslp expression, ILC2 expansion, and type 2 airway inflammation.

## MATERIAL AND METHODS

### Human subjects

We recruited 26 healthy control subjects and 57 asthma patients who were symptomatic, newly diagnosed and treatment-naïve. All subjects were Chinese and were recruited from Tongji Hospital. Subjects with asthma were diagnosed by a physician; had symptoms of episodic cough, wheeze and/or dyspnea; and had accumulated dosage of methacholine provoking a 20% fall (PD_20_) of forced expiratory volume in the first second (FEV_1_) < 2.505 mg and/or ≥12% increase in FEV_1_ following inhalation of 200μg salbutamol. We recruited subjects who had never smoked or received inhaled or oral corticosteroid or leukotriene antagonist. Healthy control subjects had no respiratory symptoms, normal spirometric value and methacholine PD_20_ ≥ 2.505 mg.

For each subject, we recorded demographic information, performed spirometry, measured fractional exhaled nitric oxide (FeNO), collected induced sputum and BALF samples. We performed bronchoscopy with endobronchial epithelial brushing. Brushing techniques was described previously [42].

### MicroRNA microarray

Total RNA from bronchial epithelial brushing samples from four type 2-low and four type 2-high asthma patients were extracted using TRIzol (Invitrogen). After passing RNA quantity measurement using the NanoDrop 1000, the samples were labeled using the miRCURY Hy3/Hy5 power labeling kit (Exiqon) and hybridized on miRCURY LNA microRNA array (7th generation, miRBase v18; Exiqon). The slides were scanned using Axon GenePix 4000B microarray scanner (Axon Instruments). Scanned images were then imported into GenePix Pro 6.0 software (Axon Instruments) for grid alignment and data extraction. We used median normalization method to obtain “normalized data”. Normalized data = (foreground - background) / median. In comparison, genes with greater than 2-fold change and statistically significant difference between two groups were considered to be differentially expressed.

### HBE cell culture and treatment

Human bronchial epithelial (HBE) cells collected from heathy donors (n = 8) by bronchial brushing technique were cultured at an air-liquid interface as previously described [43, 44]. Briefly, ten sites of the subsegmental bronchi of right middle and lower lobes were brushed. The dissociated cells were recovered by vortexing the brush in ice-cold DMEM. The cells were centrifuged, resuspended, then seeded into six-well plate coated with collagen I from rat tail (Corning) and grown in bronchial epithelial cell medium (BEpiCM; ScienCell) with supplements. Medium was changed every 48 h until cells were 90% confluent. Cells were then seeded on 1.1 cm^2^ Transwell inserts (Corning) with 0.4 μm pores. Cells were submerged for the first 7 days in BEpiCM (ScienCell) with supplements, and then the apical medium was removed to establish an air-liquid interface that was maintained for the next 14 days. The basolateral medium was changed to differentiation medium containing a 1:1 mixture of DMEM (Hyclone) and bronchial epithelial cell growth medium (BEGM; Lonza) with supplements and 50 nM all-trans retinoic acid (Sigma-Aldrich). Cells were stimulated with HDM (50 μg/ml; Greer Laboratories) and transfected with control or miR-206 mimic, control or miR-206 inhibitor (RiboBio), scrambled or CD39 siRNA, empty or CD39 cDNA expression vector. Cells were also stimulated with HDM with or without apyrase, ATPγS (Sigma-Aldrich).

### Mouse model of allergic airway inflammation

The model was established by HDM sensitization and challenge. Briefly, female C57BL/6 mice received intraperitoneal injection with 100μl of the solution of lyophilized HDM extract (0.1 mg/ml; Greer Laboratories) and Al (OH)_3_ as an adjuvant on days 0, 7 and 14, and received 40μl HDM solution (3 mg/ml) or saline intranasally on days 21, 22, and 23. MiR-206 agomir (5 nmol in 40μl saline; RiboBio) or control agomir, or miR-206 antagomir (20 nmol in 40μl saline) or control antagomir were administered intranasally on days 20 and 22. Twenty-four hours after the last HDM challenge, respiratory resistance in response to a range of concentrations of intravenous acetylcholine was measured using the forced oscillation technique with the FlexiVent system (SCIREQ) as previously described [45].

### Assessment of mouse airway inflammation

Cell counts for macrophages, eosinophils, lymphocytes, and neutrophils in bronchoalveolar lavage fluid were performed as previously described [45]. Paraffin-embedded 5-μm lung sections were stained with hematoxylin and eosin. The severity of peribronchial inflammation was scored by a blinded observer using the following features: 0, normal; 1, few cells; 2, a ring of inflammatory cells 1 cell layer deep; 3, a ring of inflammatory cells 2–4 cells deep; 4, a ring of inflammatory cells of >4 cells deep.

### Immunohistochemistry

Sections of mice lungs were stained with rabbit polyclonal CD39 (ENTPD1) antibody (Proteintech). Antibodies were detected using the Real EnVision detection system (Dako Diagnostics) according to instructions.

### PAS staining

Mice lung sections were stained with periodic acid-schiff (PAS) (Goodbio Technology) for detection of mucus. The number of PAS-staining-positive cells was counted in four random fields for each lung section at ×200 magnification.

### Quantitative polymerase chain reaction (PCR)

For quantification of hsa-miR-206 in epithelial brushings and HBE cells, mmu-miR-206 expression in mouse lungs, *CD39, IL-25, IL-33* (transcript without exons 3, 4), *TSLP* (long isoform), *CLCA1, POSTN, SERPINB2* mRNA expression in epithelial brushings, and *Cd39* mRNA expression in mouse lungs, total RNA was isolated and reverse transcribed. Quantitative PCR were performed using an ABI Prism 7500 HT Fast Real-time PCR System (Applied Biosystems). The cycle threshold (Ct) of each gene transcript was normalized to the Ct of U6, or U48 for miRNA, and to β-actin or GAPDH for mRNA, respectively. Relative gene expression was calculated by using the 2(-Delta Delta Ct) method [46]. The transcript levels of each gene are expressed as relative to the median of healthy control subjects or the mean of control group and log2 transformed. Primers for quantitative PCR were listed in Supplementary Table. The stem-loop RT primer (ssD809230234), forward primer (ssD809230926) and reverse primer (ssD089261711) for hsa-/ mmu-miR-206 were from RiboBio. Taqman primer and probe sets for *IL-25* (Hs00224471_m1), *TSLP* (long isoform; Hs01572933_m1), and *ACTB* (Hs99999903_m1) were from Applied Biosystems. The transcripts for *IL-33* without exons 3, 4 were determined by RNase H-dependent quantitative RT-PCR as reported by Gordon et al [47].

### In situ hybridization

We performed in situ hybridization of mmu-miR-206 on paraffin-embedded sections using mmu-miR-206 miRCURY LNA miRNA detection probe (Qiagen). The sequence of the probes for mmu-miR-206 was 5’-CCACACACTTCCTTACATTCCA -3’.

### Luciferase activity assay

Vector harboring wild-type, mutant CD39 3’-UTR or no 3’-UTR (control) were co-transfected with miR-206 mimic or nontargeting control into BEAS-2B cells. Luciferase activity was detected by dual luciferase reporter assay system (Promega). Normalized relative light units represent firefly luciferase activity / renilla luciferase activity.

### Western blotting

CD39 protein expression in cells was detected by monoclonal mouse-anti-human CD39 antibody (clone OTI2B10, OriGene Technologies) using Western blotting as previously described [45].

### ELISA

Human IL-25 (RayBiotech), IL-33, TSLP (R&D Systems) in supernatant from BALF and cell culture medium, and mouse Il-4, Il-5, Il-13 (R&D Systems), Il-25, Il-33 (Thermo Fisher Scientific), Tslp (R&D Systems) in BALF supernatant were measured by ELISA according to the manufacturer’s instructions. Mouse plasma IgE levels were determined by ELISA (Dakewe Biotech. All samples and standards were measured in duplicate.

### ATP measurements

To measure ATP levels in human BALF, ice-cold BALF samples were centrifuged at 4°C immediately after collection and the supernatants were stored at –80°C. Supernatant of fresh mice BALF and cell culture medium were analyzed immediately. ATP levels were measured using ATP assay kit (Beyotime Biotechnology) according to instructions.

### Flow cytometry

To analyze lung ILC2s, single cell suspensions of mouse lung tissue were incubated with cocktail of biotin-conjugated monoclonal antibodies for lineage markers [CD5, CD11b, CD45R (B220), Anti-Gr-1 (Ly-6G/C), 7-4, and Ter-119], and then mixed with Anti-Biotin MicroBeads (Miltenyi Biotec). Lineage-negative lung cells were isolated with column placed in the magnetic field of a MACS Separator (Miltenyi Biotec). Lineage-negative cells were stained with BV421-conjugated Live/Dead Fixable Dead Cell Stain (Invitrogen), PerCP/Cy5.5-conjugated CD25 (clone PC61; Biolegend), PE-conjugated CD127 (clone A7R34; Biolegend), FITC-conjugated T1/ST2 (clone DJ8; MD bioscience), APC-conjugated Sca-1 (clone D7; Biolegend). The samples were analyzed using EPICS-XL MCL flow cytometer (Beckman Coulter). Live lineage-negative CD25+CD127+T1/ST2+Sca-1+ lymphocytes were identified as ILC2s. Data were analyzed with FlowJo software (TreeStar).

### Statistics

We analyzed data using Prism version 5 (GraphPad Software) and SPSS version 19 (SPSS Inc.). For normally distributed data, we calculated means ± standard deviation (SD) and used parametric tests (unpaired Student’s *t* test or one-way ANOVA with Bonferroni’s post hoc test). For non-normally distributed data, we calculated medians (with interquartile ranges) and used non-parametric tests (Mann-Whitney test or one-way ANOVA with Bonferroni’s post hoc test). We analyzed correlation using Spearman’s rank order correlation. Values of *P* < 0.05 were considered statistically significant.

### Study approvals

Human and mouse studies were approved by the ethics committee of Tongji Hospital, Tongji Medical College, Huazhong University of Science and Technology. Participants gave informed consent.

## RESULTS

### Differentially expressed epithelial miRNAs including miR-206 between type 2-low and –high asthma

We profiled the miRNA expression of bronchial epithelial brushings from type 2-low asthmatics (*n* = 4), type 2-high asthmatics (*n* = 4) using miRNA microarray. The type 2 status of asthma was defined by the expression of the type 2 signature genes (*CLCA1, POSTN*, and *SERPINB10*) in the epithelial brushings as previously reported [3, 48]. We found that 20 miRNAs were significantly differentially expressed between the two subsets of asthma (Figure 1A). The data are available at GEO (http://www.ncbi.nlm.nih.gov/geo/, accession number GSE142237). Of note, miR-206 was the most highly expressed miRNA in type 2-high asthma relative to type 2-low asthma. Several other differentially expressed miRNAs including miR-31-5p, miR-146a-5p, miR-146b-5p, miR-221-3p have been implicated in asthma pathogenesis [49-51]. Epithelial miR-221-3p expression was shown to be associated with airway eosinophilia and the expression of type 2 signature genes in asthma patients in our previous study [52].

**Figure 1.**
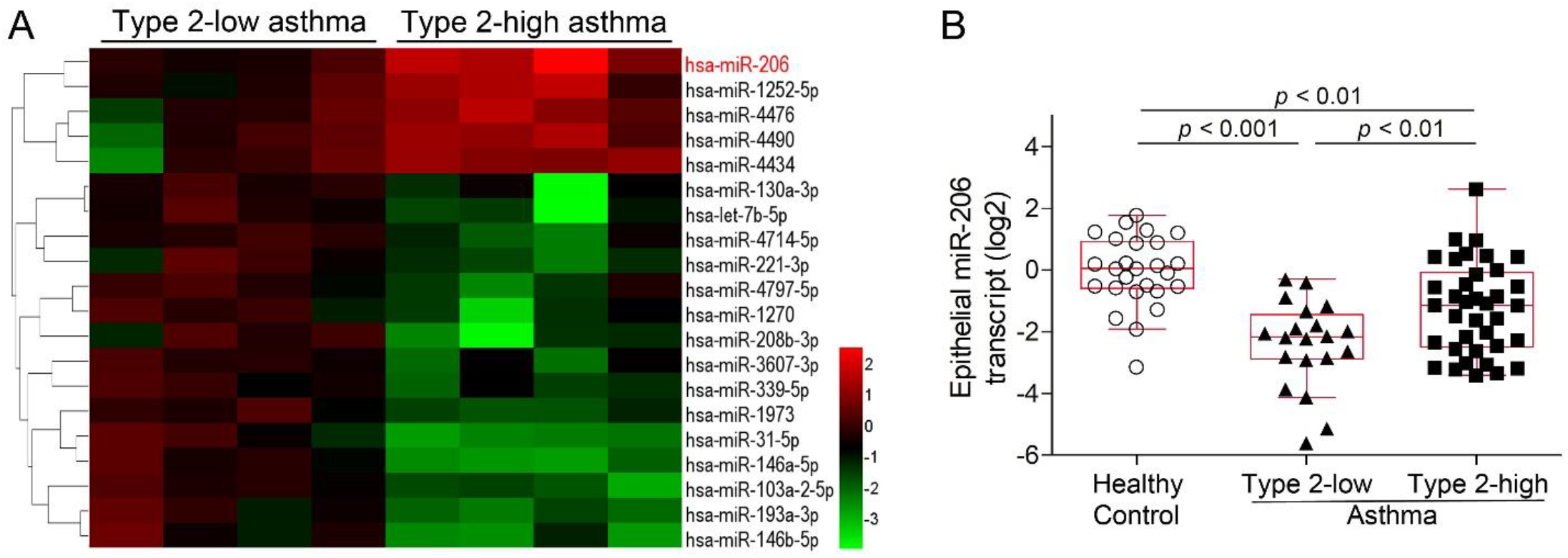
Epithelial miRNA expression profiling reveals differentially expressed miRNAs including miR-206 between type 2-low and –high asthma. (A) Twenty differentially expressed epithelial miRNAs between subjects with type 2-low asthma (n = 4) and type 2-high asthma (n = 4) were identified using microarrays. Each column represents data from one subject. Colors represent fold change relative to the mean value of type 2-low asthma. (B) miR-206 transcript levels in bronchial brushings from type 2-low asthma patients (n = 20), type 2-high asthma patients (n = 37) and healthy controls (n = 26) were determined by quantitative PCR. The transcript levels are expressed as relative to the median value of healthy controls and log2 transformed. One-way ANOVA with Bonferroni’s post hoc test was performed.

We next examined the expression of miR-206 in a cohort including type 2-low asthmatics (*n* = 20), type 2-high asthmatics (*n* = 37), and healthy controls (*n* = 26) using quantitative PCR. Compared with type 2-low asthmatics, type 2-high asthmatics had lower methacholine PD_20_, higher eosinophil counts in induced sputum and blood, higher fractional exhaled nitric oxide (FeNO) and serum IgE levels (Table 1). Consistent with the microarray data, epithelial miR-206 expression was higher in type 2-high asthma relative to type 2-low asthma. However, compared to healthy controls, epithelial miR-206 transcript levels were decreased in both type 2-low and type 2-high asthma (Figure 1B). Our data suggest that epithelial miR-206 expression is downregulated in asthma and associates with airway type 2 inflammation.

**Table 1.**
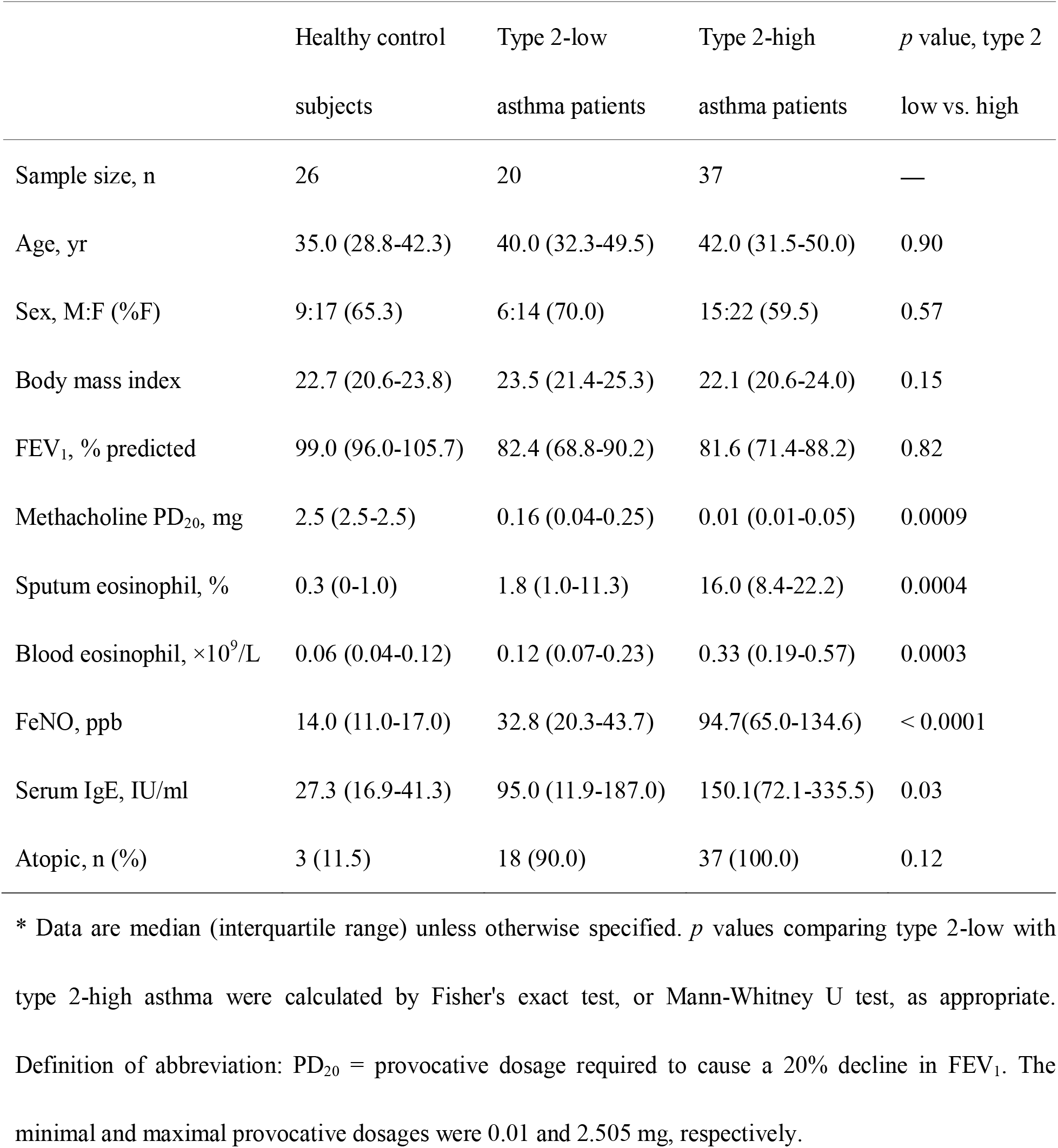
Subjects Characteristics*.

|  | Healthy control | Type 2-low | Type 2-high | *p* value, type 2 |
| --- | --- | --- | --- | --- |
|  | subjects | asthma patients | asthma patients | low vs. high |
| Sample size, n | 26 | 20 | 37 | — |
| Age, yr | 35.0 (28.8-42.3) | 40.0 (32.3-49.5) | 42.0 (31.5-50.0) | 0.90 |
| Sex, M:F (%F) | 9:17 (65.3) | 6:14 (70.0) | 15:22 (59.5) | 0.57 |
| Body mass index | 22.7 (20.6-23.8) | 23.5 (21.4-25.3) | 22.1 (20.6-24.0) | 0.15 |
| FEV<sub>1</sub>, % predicted | 99.0 (96.0-105.7) | 82.4 (68.8-90.2) | 81.6 (71.4-88.2) | 0.82 |
| Methacholine PD<sub>20</sub>, mg | 2.5 (2.5-2.5) | 0.16 (0.04-0.25) | 0.01 (0.01-0.05) | 0.0009 |
| Sputum eosinophil, % | 0.3 (0-1.0) | 1.8 (1.0-11.3) | 16.0 (8.4-22.2) | 0.0004 |
| Blood eosinophil, ×10<sup>9</sup>/L | 0.06 (0.04-0.12) | 0.12 (0.07-0.23) | 0.33 (0.19-0.57) | 0.0003 |
| FeNO, ppb | 14.0 (11.0-17.0) | 32.8 (20.3-43.7) | 94.7(65.0-134.6) | < 0.0001 |
| Serum IgE, IU/ml | 27.3 (16.9-41.3) | 95.0 (11.9-187.0) | 150.1(72.1-335.5) | 0.03 |
| Atopic, n (%) | 3 (11.5) | 18 (90.0) | 37 (100.0) | 0.12 |

\* Data are median (interquartile range) unless otherwise specified. *p* values comparing type 2-low with type 2-high asthma were calculated by Fisher's exact test, or Mann-Whitney U test, as appropriate. Definition of abbreviation: PD<sub>20</sub> = provocative dosage required to cause a 20% decline in FEV<sub>1</sub>. The minimal and maximal provocative dosages were 0.01 and 2.505 mg, respectively.

### The expression of CD39, a target of miR-206, is upregulated in airway epithelial cells in asthma

We predicted the candidate target genes of miR-206 by using online algorithms (DIANA-microT, miRanda, miRWalk, PicTar and TargetScan). CD39, the ectoenzyme catalyzing the degradation of extracellular ATP and ADP, is one of the candidate targets. The 3’-untranslated region (UTR) of CD39 contains nucleotides matching the seed sequence of hsa-miR-206 (Figure 2A). Transfection of miR-206 mimic decreased the luciferase activity when co-transfected with the vector harboring wild-type CD39 3’-UTR, but had no effect on the luciferase activity when co-transfected with the vector containing mutant 3’-UTR or empty vector (Figure 2B). This indicates that miR-206 may directly act on the 3’-UTR of CD39 mRNA. Further, inhibition of miR-206 expression enhanced CD39 mRNA and protein expression, whereas overexpression of miR-206 suppressed CD39 expression (Figure 2C-E). These data indicate that CD39 is a target gene of miR-206.

**Figure 2.**
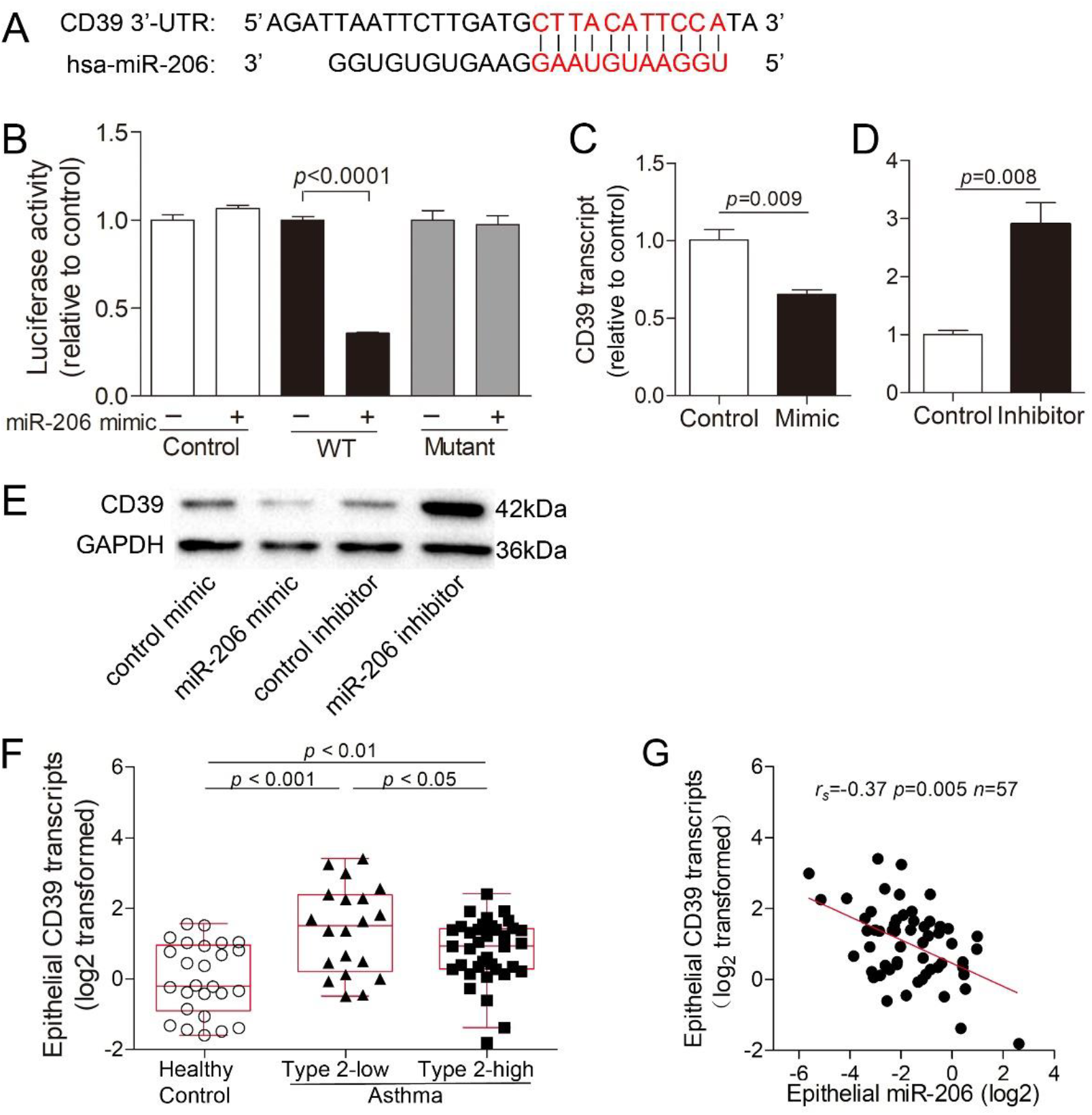
The expression of CD39, a target of miR-206, is increased in airway epithelial cells in human asthma. (A) The 3’-UTR of CD39 contains region that matches the seed sequence of hsa-miR-206. (B) 3’-UTR luciferase report assay with vector harboring wild-type (WT), mutant CD39 3’-UTR or no 3’-UTR (control) co-transfected with miR-206 mimic or non-targeting control. Luciferase activity was measured by dual-luciferase reporter assay system. The firefly luciferase activity was normalized to renilla luciferase activity. n = 4 per group. (C, D) CD39 transcript levels in BEAS-2B cells after transfection with miR-206 mimic (*C*) or inhibitor (*D*) were determined by quantitative PCR. The transcript levels are expressed as relative to the mean value of controls (two-tailed Student’s *t* test). n = 4 per group. Data are mean ± SD. (E) CD39 protein expression in BEAS-2B cells after transfection with miR-206 mimic and inhibitor were determined by Western blotting. (F) CD39 transcript levels in bronchial brushings from type 2-low asthma patients (n = 20), type 2-high asthma patients (n=37) and healthy controls (n = 26) were determined by quantitative PCR. The transcript levels are expressed as relative to the median value of controls and log2 transformed (one-way ANOVA with Bonferroni’s post hoc test). (G) Spearman’s rank order correlation assay between epithelial CD39 and miR-206 transcript levels in all asthma patients (n = 57).

In human asthma, we found that CD39 transcript levels were significantly increased in bronchial brushings from type 2-low and -high asthma patients compared to controls. Moreover, epithelial CD39 expression was lower in type 2-high asthma relative to type 2-low asthma (Figure 2F). In support of CD39 as a target of miR-206, epithelial CD39 transcript levels negatively correlated with epithelial miR-206 expression (Figure 2G) in asthma patients.

### Extracellular ATP accumulation induces miR-206 downregulation and CD39 upregulation in human bronchial epithelial cells

It was reported that BALF ATP concentration was increased in asthma patients after allergen provocation [28]. In BALF from our symptomatic and treatment-naïve asthma patients, we measured ATP using luciferase bioluminescence. BALF ATP levels were markedly increased in both type 2-low and –high asthmatics compared to controls (Figure 3A). Moreover, BALF ATP levels were higher in type 2-high asthma relative to type 2-low asthma. This suggests that extracellular ATP is accumulated in the airways of symptomatic asthma patients.

**Figure 3.**
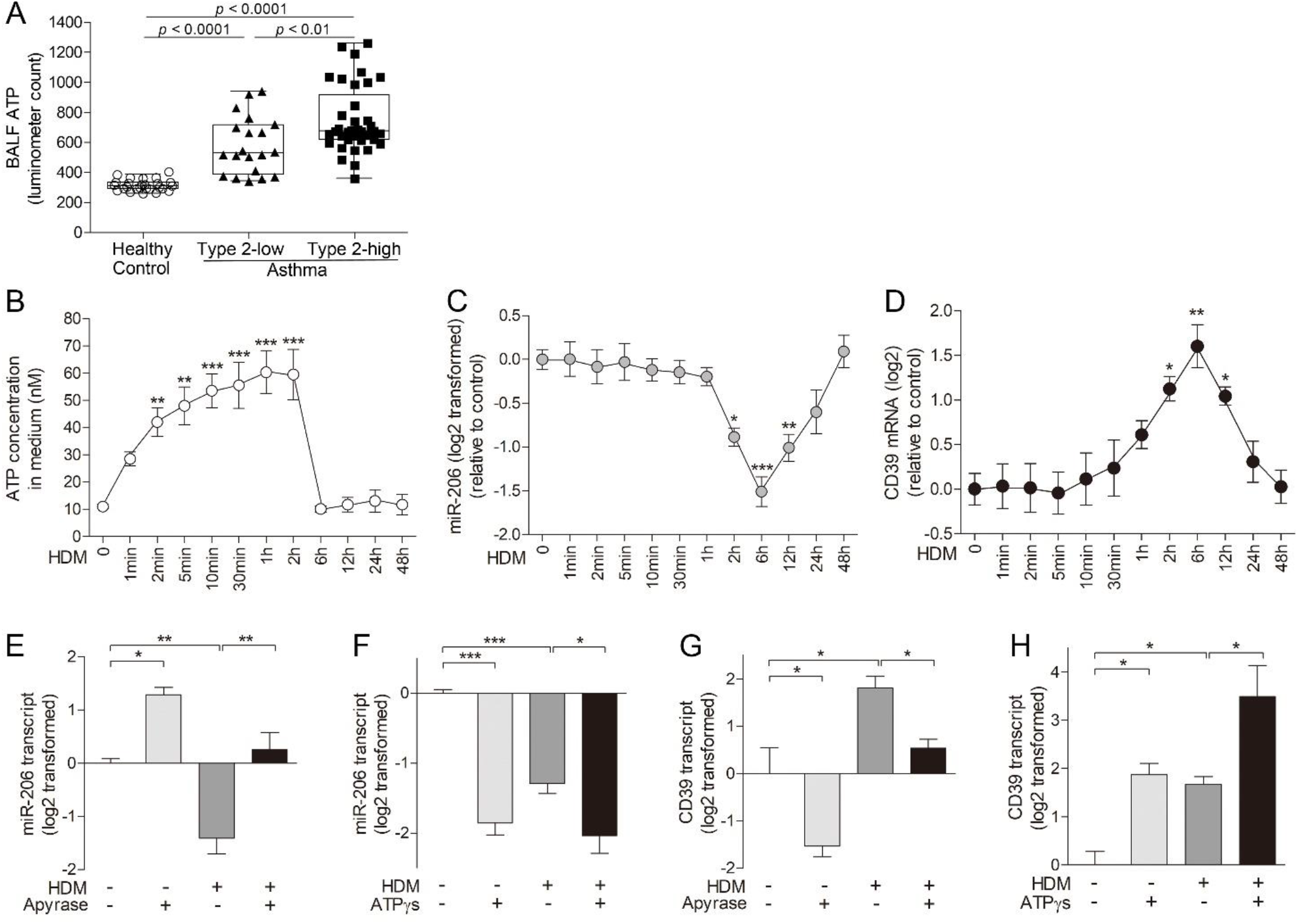
Acute extracellular ATP accumulation is responsible for allergen-induced miR-206 downregulation and CD39 upregulation in bronchial epithelial cells. (A) ATP levels in BALF from type 2-low asthma patients (n = 20), type 2-high asthma patients (n = 37), and healthy controls (n = 26) were determined by luciferase bioluminescence. BALF ATP levels were expressed as luminometer counts (one-way ANOVA with Bonferroni’s post hoc test). (B) ATP concentration in culture medium collected at indicated time points after HDM stimulation was measured by luciferase bioluminescence. (C, D) Transcript levels of miR-206 (*C*) and CD39 (*D*) in HBE cells harvested at indicated time points after HDM stimulation were determined by quantitative PCR. n = 4-6 per time point combined from 2 experiments using HBE cells from 2 healthy donors. Data are mean ± SD. \**P*<0.05; \*\**P*<0.01; \*\*\**P*<0.001 (one-way ANOVA with Bonferroni’s post hoc test). (E-F) MiR-206 transcript levels in HBE cells pretreated with apyrase or saline for 2 h before adding HDM and stimulation for 6 h (*E*), and treated with ATPγS or saline with or without HDM for 6h (*F*). (G, H) CD39 transcript levels in HBE cells pretreated with apyrase or saline for 2 h before adding HDM and stimulation for 6 h (*G*), and treated with ATPγS or saline with or without HDM for 6h (*H*). The transcript levels are expressed as relative to the mean value of control group and log2 transformed. n = 4-6 wells per group combined from 2 experiments using HBE cells from 2 healthy donors. Data are mean ± SD. \**P*<0.05; \*\**P*<0.01; \*\*\**P*<0.001 (one-way ANOVA with Bonferroni’s post hoc test).

Extracellular ATP accumulation upregulates CD39 expression in airway epithelial cells, which catalyzes the degradation of excessive extracellular ATP to maintain the homeostasis of microenvironment [53]. To determine whether miR-206-CD39 axis responds to extracellular ATP accumulation, we performed air-liquid interface culture of human bronchial epithelial (HBE) cells from healthy donors. House dust mite (HDM), the most clinically relevant allergen, rapidly increased ATP concentration in culture medium within 2 min. The ATP concentration peaked at 1-2 h, and declined to baseline by 6h (Figure 3B). HDM exposure decreased miR-206 expression from 2 h to 12 h, with a maximum inhibition at 6 h (Figure 3C), and increased CD39 mRNA from 2 h to 12 h, peaking at 6 h (Figure 3D). Furthermore, elimination of extracellular ATP by pretreatment with apyrase suppressed HDM-induced miR-206 downregulation and CD39 upregulation at 6 h in HBE cells (Figure 3E and G). Exogenous ATP analog, ATPγS, directly decreased miR-206 expression and increased CD39 expression, and intensified HDM-induced miR-206 downregulation and CD39 upregulation (Figure 3F and H). Together, our data suggest that allergen-induced acute accumulation of extracellular ATP downregulates miR-206 and upregulates CD39 expression in airway epithelial cells. This may represent a protective mechanism to eliminate excessive extracellular ATP.

### Higher ATP levels associate with elevated IL-25 and TSLP expression in type 2-high asthma

Allergen stimulates the release of ATP as an alarmin from airway epithelial cells to induce the expression of IL-33 [34]. We next examined the airway IL-25, IL-33 and TSLP expression in bronchial epithelial brushings and BALF using quantitative PCR and ELISA, respectively. Epithelial *IL25* transcript levels and BALF IL-25 protein levels were significantly higher in type 2-high asthma when compared with type 2-low asthma and controls (Figure 4A, 4D). There were multiple splice variants of *IL-33* transcript, and the protein encoded by the *IL33* transcript without exons 3 and 4 is secreted as active cytokine [47]. We examined the expression of *IL33* transcript without exons 3 and 4 in bronchial epithelial brushings using RNase H-dependent quantitative PCR as previously reported [47]. However, there were no significant difference in this *IL33* transcript among the two asthma subsets and controls. Neither did we detect significant difference in IL-33 protein levels in BALF (Figure 4B, 4E). TSLP has short and long isoforms, and the long isoform is induced during inflammation [54]. We found that the long *TSLP* transcripts and BALF TSLP protein levels were higher in type 2-high asthma compared to type 2-low asthma and control subjects (Figure 4C, 4F). Our data suggest that airway expression of IL-25, TSLP whereas not IL-33 is elevated in type 2-high asthma.

**Figure 4.**
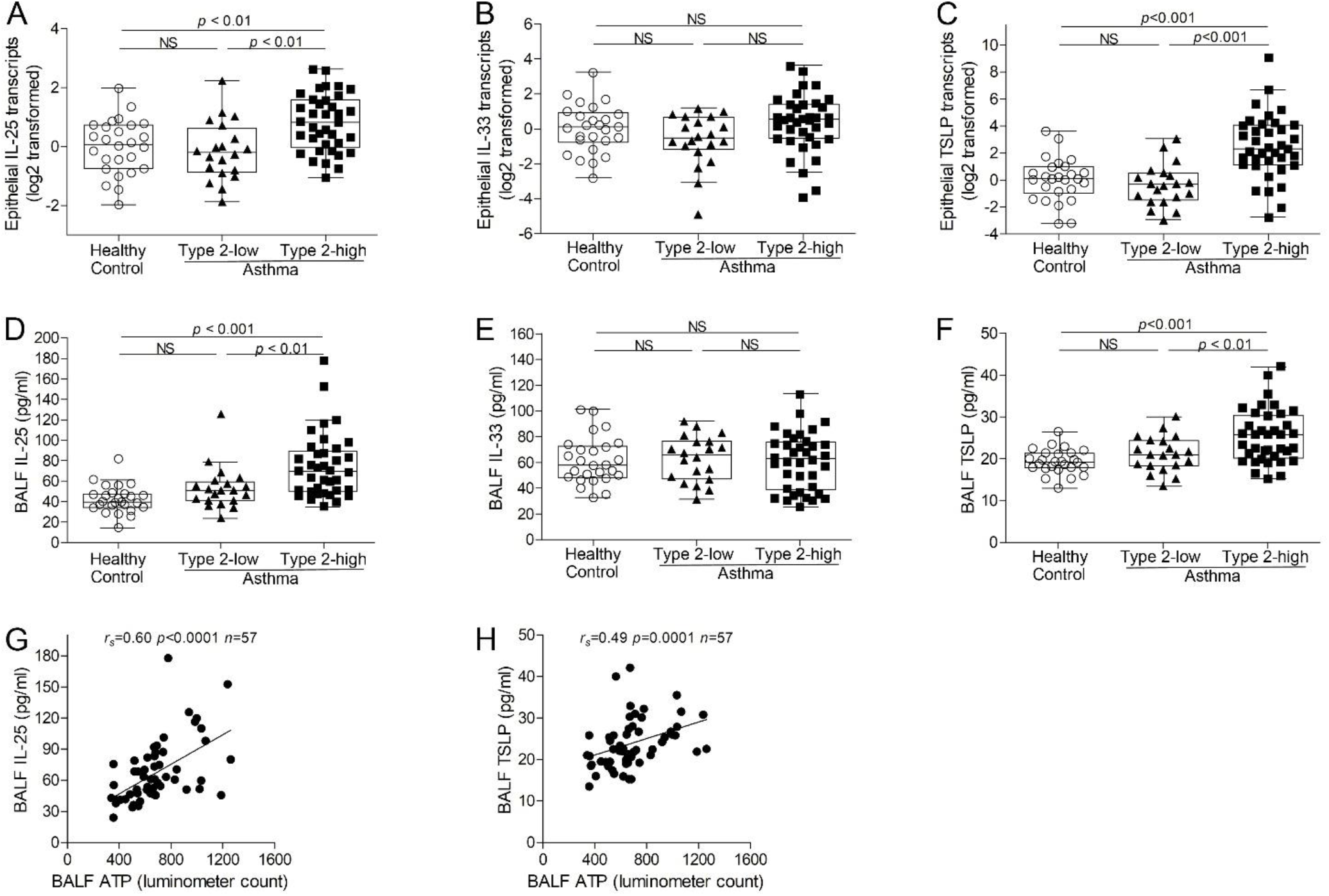
Airway IL-25 and TSLP expression is elevated in type 2-high asthma, and correlates with BALF ATP levels in asthma. (A-C) The transcripts of *IL-25* (*A*), *IL-33* without exons 3, 4 (*B*), and long isoform of *TSLP* (*C*) in bronchial epithelial brushings from type 2-low asthma (n = 20), type 2-high asthma (n = 37) and healthy controls (n = 26) were determined using quantitative PCR with Taqman primers and probes. For detection of *IL-33* transcript without exons 3 and 4, RNase H-dependent quantitative PCR was performed. The transcript levels are expressed as relative to the median value of healthy controls and log2 transformed. (D-F) IL-25 (*D*), IL-33 (*E*) and TSLP (*F*) protein levels in BALF from type 2-low asthma (n = 20), type 2-high asthma (n = 37) and healthy controls (n = 26) were determined using ELISA. One-way ANOVA with Bonferroni’s post hoc test was performed. (G-H) Spearman’s rank order correlation assays between BALF ATP levels and BALF IL-25 protein levels (*G*), and TSLP protein levels (*H*).

Importantly, BALF ATP levels were strongly correlated with BALF IL-25 and TSLP protein levels (Figure 4G-H). This indicates that the more prominent accumulation of airway ATP may be responsible for elevated IL-25 and TSLP expression in type 2-high asthma.

### Extracellular ATP is essential for allergen-induced IL-25 and TSLP expression in human bronchial epithelial cells

We next examined the role of extracellular ATP in HDM-induced IL-25, IL-33 and TSLP expression in HBE cells cultured on an air-liquid interface. We found that HDM stimulation increased *IL-25* mRNA expression in HBE cells and IL-25 protein levels in basal-lateral medium, peaking at 2 h and 6 h, respectively (Figure 5A and D). HDM also increased TSLP (the long transcript variant) mRNA and protein expression, peaking at 1 h and 6 h, respectively (Figure 5C and F). However, HDM stimulation did not alter the expression of IL-33 (the transcript without exons 3, 4) mRNA or protein (Figure 5B and E). Elimination of extracellular ATP by CD39 overexpression or by using apyrase suppressed HDM-induced IL-25 and TSLP protein expression at 6 h (Figure 5G-J). In contrast, enhancing extracellular ATP by CD39 knockdown or by adding ATPγS further increased HDM-induced IL-25 and TSLP protein expression (Figure 5K-N). These data suggest that HDM-induced acute accumulation of extracellular ATP is required for IL-25 and TSLP upregulation in airway epithelial cells.

**Figure 5.**
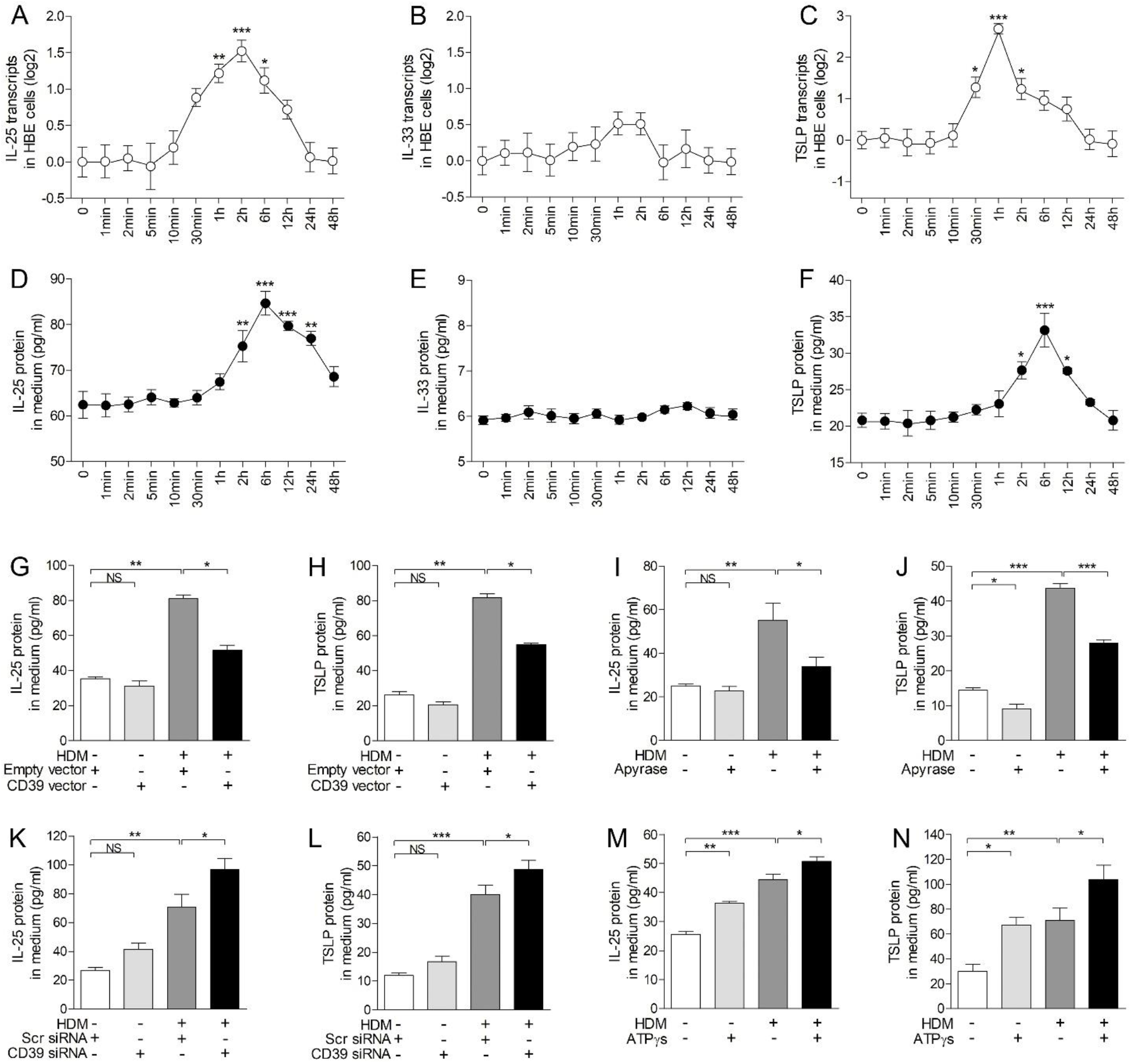
Extracellular ATP is required and sufficient for IL-25 and TSLP expression in bronchial epithelial cells. (A-C) The transcripts of *IL-25* (A), *IL-33* without exons 3, 4 (*B*), and the long isoform of TSLP (C) in HBE cells harvested at indicated time points after HDM stimulation were determined by quantitative PCR. (D-F) IL-25 (*D*), IL-33 (*E*) and TSLP (*F*) protein levels in culture medium collected at indicated time points after HDM stimulation were determined by ELISA. \**P*<0.05; \*\**P*<0.01; \*\*\**P*<0.001 (one-way ANOVA with Bonferroni’s post hoc test). n = 5-6 wells per time points combined from 2 experiments using HBE cells from 2 healthy donors. (G, H) IL-25 (G) and TSLP (*H*) protein levels in culture medium after transfection with empty or CD39 expression vector and stimulated with or without HDM for 6 h were determined by ELISA. (I, J) IL-25 (*I*) and TSLP (*J*) protein levels in culture medium after pretreatment with apyrase or saline and stimulated with or without HDM for 6 h were determined by ELISA. (K, L) IL-25 (*K*) and TSLP (*L*) protein levels in culture medium after transfection with scrambled or CD39 siRNA and stimulated with or without HDM for 6 h were determined by ELISA. (M, N) IL-25 (*M*) and TSLP (*N*) protein levels in culture medium after treatment with ATPγS or saline and with or without HDM for 6h were determined by ELISA. n = 4 wells per group. Data are mean ± SD. \**P*<0.05; \*\**P*<0.01; \*\*\**P*<0.001 (one-way ANOVA with Bonferroni’s post hoc test). n = 4-6 wells per group combined from 2 experiments using HBE cells from 2 healthy donors.

### MiR-206 regulates allergen-induced IL-25 and TSLP expression in bronchial epithelial cells via targeting CD39-extracellular ATP axis

We next examined whether miR-206 regulates IL-25 and TSLP expression via targeting CD39-extracellular ATP axis in HDM-stimulated HBE cells. Inhibition of miR-206 expression by transfection with miR-206 inhibitor increased baseline and HDM-induced CD39 expression. Importantly, inhibition of miR-206 expression decreased the ATP concentration, and suppressed HDM-induced IL-25 and TSLP protein expression in the medium (Figure 6A-D). In contrast, miR-206 overexpression by transfection with miR-206 mimic suppressed baseline and HDM-induced CD39 expression, enhanced extracellular ATP, and further enhanced HDM-induced IL-25 and TSLP protein expression (Figure 6E-H). Our data indicates that miR-206 regulates epithelial IL-25 and TSLP expression by targeting CD39-extracellular ATP axis.

**Figure 6.**
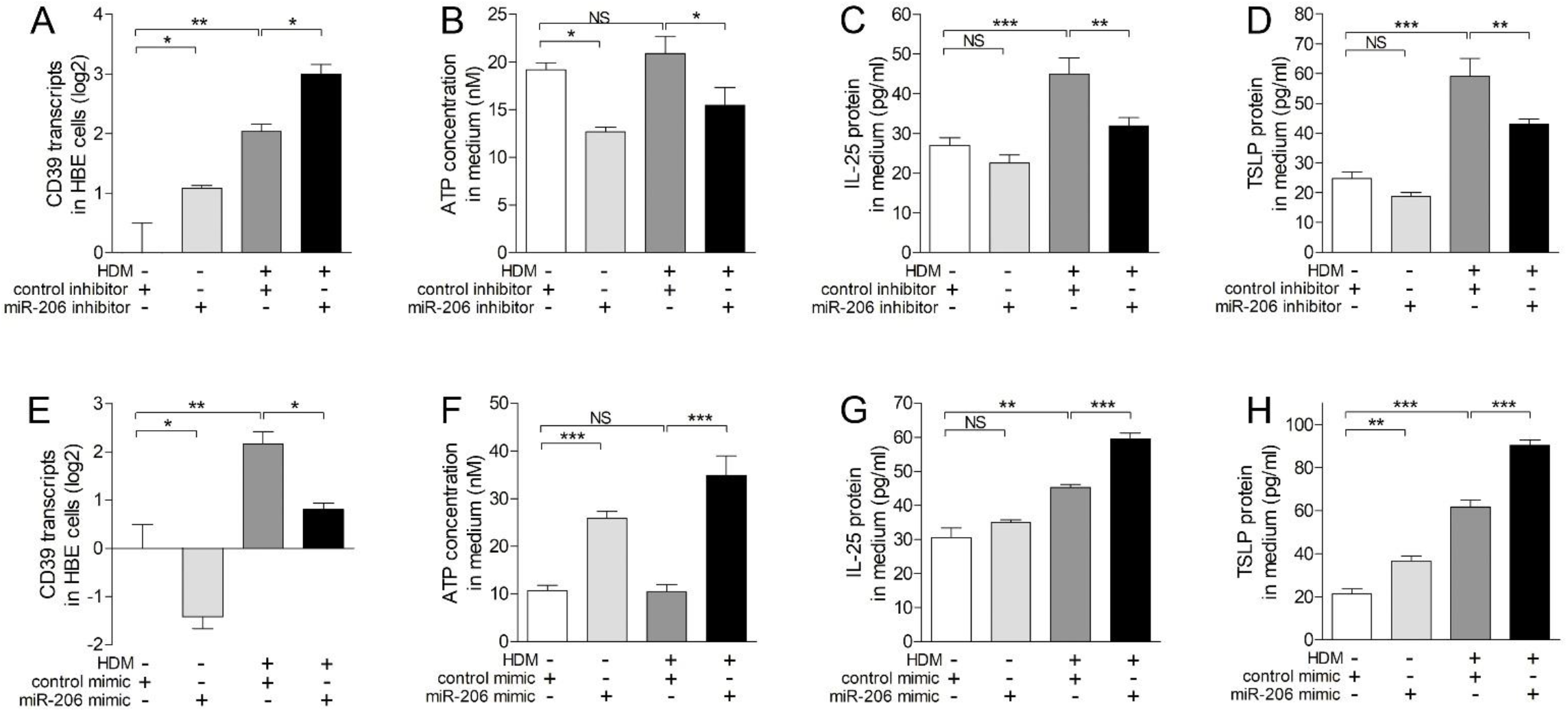
MiR-206 regulates allergen-induced IL-25 and TSLP expression via targeting CD39 - extracellular ATP axis in bronchial epithelial cells. (A) CD39 transcript levels in HBE cells transfected with control or miR-206 inhibitor and stimulated with or without HDM for 6 h were determined by quantitative PCR. (B) ATP concentration in culture medium after transfection with control or miR-206 inhibitor and stimulation with or without HDM for 6 h were measured by luciferase bioluminescence. (C, D) IL-25 (*C*), and TSLP (*D*) protein levels in culture medium after transfection with control or miR-206 inhibitor and stimulation with or without HDM for 6 h were determined by ELISA. (E) CD39 transcript levels in HBE cells transfected with control or miR-206 mimic and stimulated with or without HDM for 6 h were determined by quantitative PCR. (F) ATP concentration in culture medium after transfection with control or miR-206 mimic and stimulation with or without HDM for 6 h were measured by luciferase bioluminescence. (G, H) IL-25 (*G*), and TSLP (*H*) protein levels in culture medium after transfection with control or miR-206 mimic and stimulation with or without HDM for 6 h were determined by ELISA. n = 4 wells per group. Data are mean ± SD. \**P*<0.05; \*\**P*<0.01; \*\*\**P*<0.001 (one-way ANOVA with Bonferroni’s post hoc test). n = 5-6 wells per group combined from 2 experiments using HBE cells from 2 healthy donors.

### Airway epithelial miR-206 regulates CD39 expression, BALF ATP concentration in a murine model of allergic airway disease

We investigated the role of epithelial miR-206 in a murine model of allergic airway inflammation of C57BL/6 mice sensitized and challenged with HDM. Using quantitative PCR, in situ hybridization, immunohistochemistry and luciferase bioluminescence, we found that HDM challenge decreased epithelial miR-206 expression, increased CD39 expression, and markedly enhanced BALF ATP levels compared to control mice. Inhibition of airway miR-206 expression by intranasal administration of miR-206 antagomir further enhanced HDM-induced CD39 expression whereas suppressed HDM-induced BALF ATP accumulation (Figure 7A-D). In contrast, airway overexpression of miR-206 by intranasal administration of miR-206 agomir significantly suppressed HDM-induced CD39 expression, and further enhanced HDM-induced increase of ATP concentrations (Figure 7E-I). These data suggest that epithelial miR-206 targets CD39-extracellular ATP axis in the airway of a murine model of allergic airway disease.

**Figure 7.**
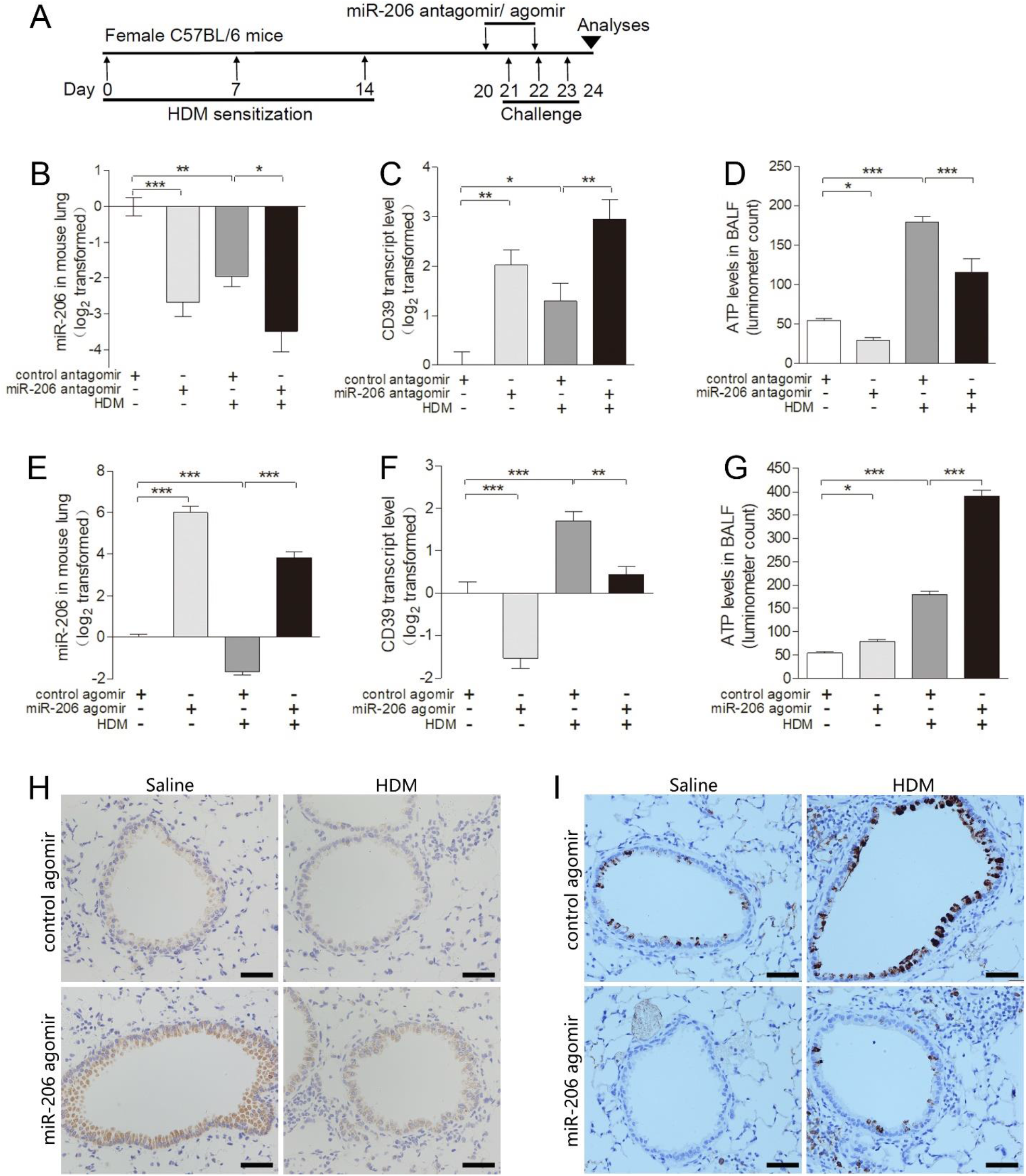
Epithelial miR-206 targets CD39-extracellular ATP axis in a murine model of allergic airway inflammation. (A) Experimental schedule as described in Methods. (B-D) MiR-206 (*B*) and CD39 (*C*) transcript levels in lungs, and ATP levels in BALF (*D*) were determined by quantitative PCR and luciferase bioluminescence, respectively, in mice intranasally administered with control or miR-206 antagomir and challenged with HDM or saline. (E-G) MiR-206 (*E*) and CD39 (*F*) transcript levels in lungs, and ATP levels in BALF (*G*) were determined by quantitative PCR and luciferase bioluminescence, respectively, in mice intranasally administered with control or miR-206 agomir and challenged with HDM or saline. The transcript level is expressed as relative to the mean value of control group and log2 transformed. BALF ATP levels were expressed as luminometer counts. n = 6 - 10 mice per group combined from 2 independent experiments. Data are mean ± SD. \**P* < 0.05; \*\**P* < 0.01; \*\*\**P* < 0.001 (one-way ANOVA with Bonferroni’s post hoc test). (H, I) Representative images of *in situ* hybridization of miR-206 (*H*), and immunohistochemistry of CD39 (*I*) in lung sections of mice intranasally administered with control or miR-206 agomir and challenged with HDM or saline. Scale bar = 50 μm.

### Airway miR-206 antagonism suppresses HDM-induced AHR, airway eosinophilia, mucus overproduction and type 2 cytokines expression in mice

In the murine model of allergic airway disease, HDM sensitization and challenge increased airway resistance to methacholine, induced the infiltration of inflammatory cells around airways, increased airway mucus-producing cells and plasma IgE levels. Inhibition of airway miR-206 expression by transfection with miR-206 antagomir significantly suppressed HDM-induced AHR, airway eosinophilic inflammation and mucus overproduction (Figure 8A-F). In contrast, airway miR-206 overexpression by transfection with miR-206 agomir further enhanced HDM-induced AHR, airway eosinophilia and mucus overproduction (Supplementary Figure A-F). Moreover, airway miR-206 antagonism suppressed HDM-induced expression of type 2 cytokines including Il-4, Il-5 and Il-13, and decreased plasma IgE levels (Figure 8G-J), whereas miR-206 overexpression further enhanced HDM-induced expression of type 2 cytokines in mice lungs and plasma IgE levels (Supplementary Figure G-J). Our data indicates that epithelial miR-206 play an essential role in allergic airway disease by regulating type 2 immune response.

**Figure 8.**
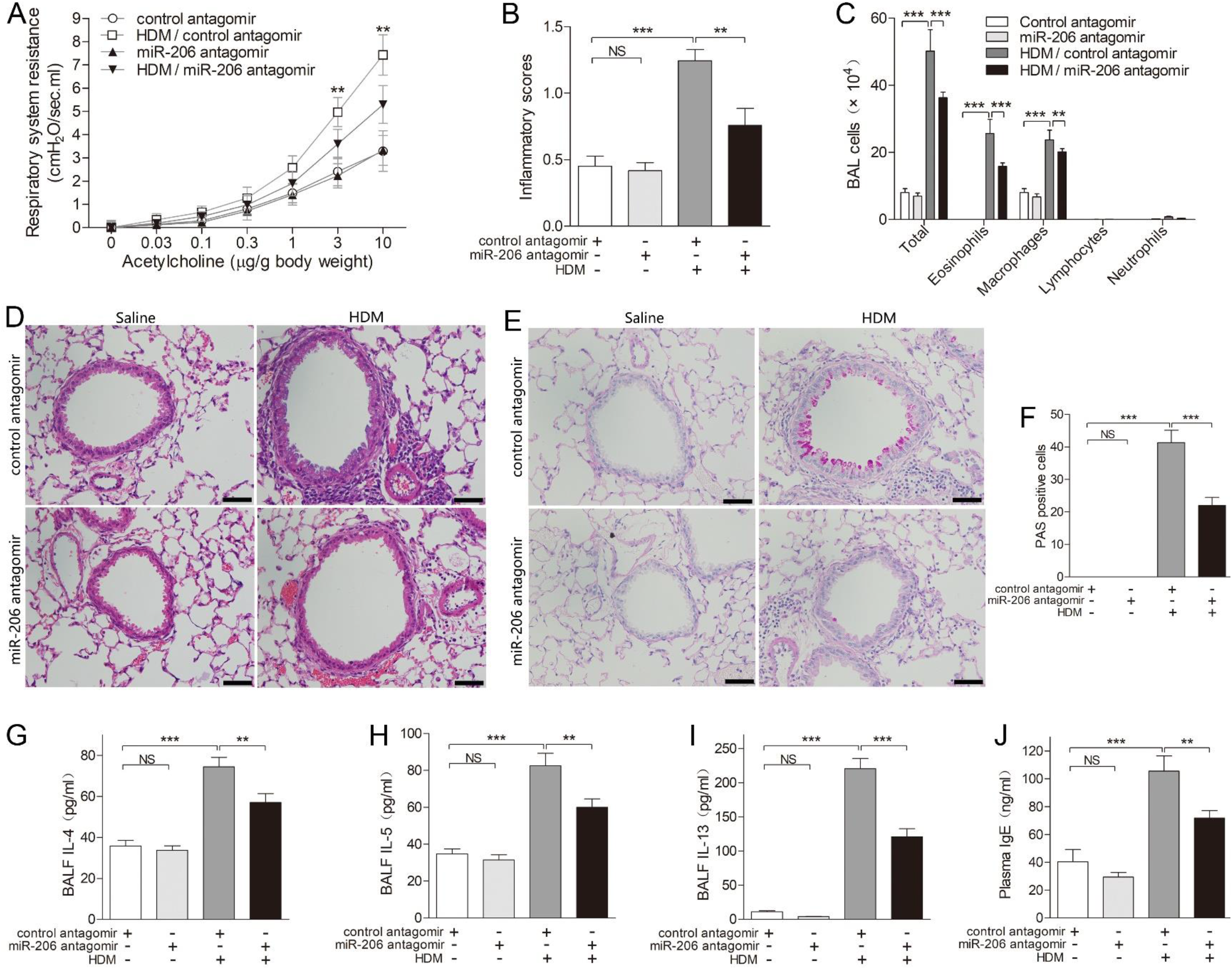
Airway miR-206 antagonism suppresses HDM-induced AHR, airway inflammation, mucus overproduction and type 2 response in mice. (A) Respiratory resistance in response to different concentration of intravenous acetycholine at 24 h after the last HDM or saline challenge in mice intranasally administered with control or miR-206 antagomir. (B) Inflammatory scores of lung sections from mice intranasally administered with control or miR-206 antagomir and challenged with HDM or saline were calculated as described in Methods. (C) Counts for macrophages, eosinophils, lymphocytes and neutrophils in BALF. (D) H&E staining of representative lung sections. (E) PAS staining for mucus in representative lung sections. (F) The numbers of PAS-staining-positive cells were counted in four random fields for each lung section at ×200 magnification. (G-I) The protein levels of IL-4 (*G*), IL-5 (*H*), IL-13 (*I*) in BALF were determined by ELISA. (J) Plasma IgE levels in peripheral blood were determined by ELISA. n = 6-10 mice per group combined from 2 independent experiments. Data are mean ± SD. \**P*<0.05; \*\**P*<0.01; \*\*\**P*<0.001 (one-way ANOVA with Bonferroni’s post hoc test). Scale bar = 50 μm.

### Airway miR-206 antagonism suppresses Il-25, Il-33, Tslp expression and ILC2s expansion in mice

Since ILC2s play a pivotal role in type 2 response upon activation by IL-25 IL-33 and TSLP, we further examined the effect of airway miR-206 manipulation on Il-25, Il-33, Tslp expression and ILC2s expansion in mice. HDM sensitization and challenge increased Il-25, Il-33 and Tslp protein levels in BALF (Figure 9A-F). Meanwhile, HDM sensitization and challenge significantly increased the number of ILC2s (Lin^-^CD25^+^CD127^+^ST2^+^Sca-1^+^, gating strategy in Figure 9G) in single-cell suspension of lungs by flow cytometric analysis (Figure 9H-K). Airway miR-206 antagonism suppressed HDM-induced Il-25, Il-33, and Tslp expression (Figure 9A-C) and ILC2s expansion (Figure 9H and J), whereas miR-206 overexpression further enhanced HDM-induced Il-25, Il-33 and Tslp expression (Figure 9D-F) and ILC2s expansion in mice lungs (Figure 9I and K). This suggests that epithelial miR-206 regulates HDM-induced Il-25, Il-33 and Tslp expression and ILC2s expansion in mouse models.

**Figure 9.**
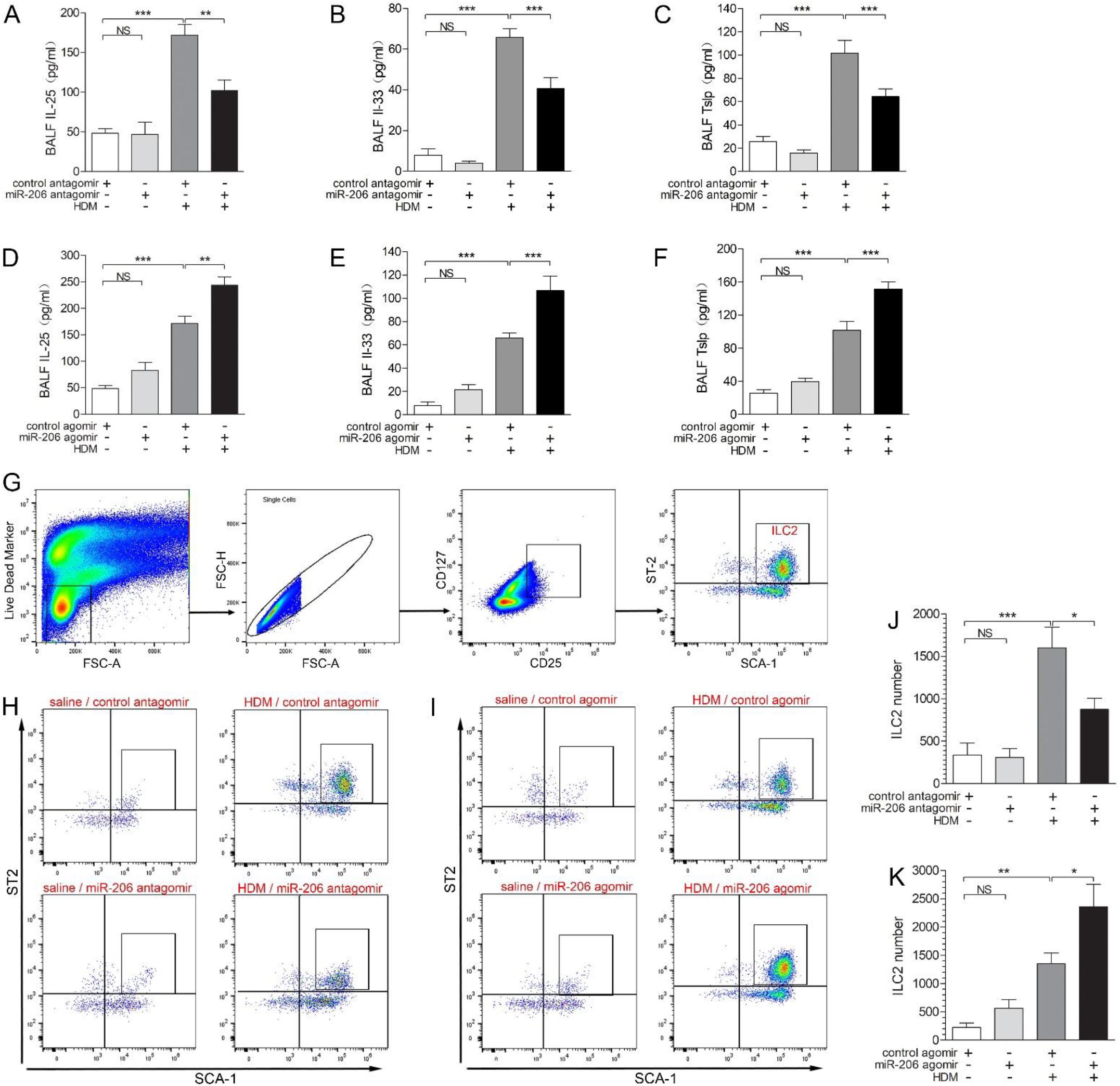
Perturbation of airway miR-206 expression alters HDM-induced Il-25, Il-33, Tslp expression and ILC2 expansion in mice lung. (A-C) Il-25 (*A*), Il-33 (*B*) and Tslp (*C*) protein levels in BALF were determined by ELISA in mice intranasally administered with control or miR-206 antagomir and challenged with HDM or saline. (D-F) Il-25 (*D*), Il-33 (*E*) and Tslp protein levels (*F*) in BALF were determined by ELISA in mice intranasally administered with control or miR-206 agomir and challenged with HDM or saline. n = 6-10 mice per group combined from 2 independent experiments. (G) Single cell suspensions of mouse lung tissue were incubated with cocktail of biotin-conjugated antibodies for lineage markers and mixed with Anti-Biotin MicroBeads to isolate lineage-negative lung cells. ILC2s in mice lungs were enumerated via flow cytometry analysis with lineage-negative lung cells using the following gating strategy: live, single, CD25^+^ CD127^+^ ST2^+^ Sca-1^+^ cells. (H, J) Representative flow cytometric plots (*H*) and numbers of ILC2s (*J*) from lungs of mice intranasally administered with control or miR-206 antagomir and challenged with HDM or saline. (I, K) Representative flow cytometric plots (*I*) and numbers of ILC2s (*K*) from lungs of mice intranasally administered with control or miR-206 agomir and challenged with HDM or saline. n = 4 -6 per group combined from 2 experiments. Data are mean ± SD. \**P*<0.05; \*\**P*<0.01; \*\*\**P*<0.001 (one-way ANOVA with Bonferroni’s post hoc test).

## DISCUSSION

In the present study, we reported that epithelial miR-206 was differentially expressed between type 2-low and -high asthma patients who were symptomatic and treatment-naïve. Type 2-high asthma patients had higher miR-206 expression, lower epithelial CD39 expression, elevated BALF ATP levels, and higher epithelial IL-25, TSLP expression as compared with type 2-low asthma. Of note, BALF ATP levels strongly correlated with airway IL-25 and TSLP expression in asthma patients. The associations between these measurements were functionally validated in primary culture of HBE cells, and in a murine model of allergic airway inflammation.

MiRNAs play essential roles in the pathogenesis of asthma [39, 40, 55, 56]. So far, the differentially expressed epithelial miRNAs between type 2-low and high asthma remains unknown. We identified miR-206 as the most highly miRNA in type 2-high asthma relative to type 2-low asthma. However, compared to control subjects, miR-206 expression was downregulated in both asthma subsets. MiR-206 expression was also decreased in cultured HBE cells exposed to HDM and in the airways of mouse sensitized and challenged with HDM. Our findings are consistent with the report that miR-206 expression was decreased in airway wall of a mouse model of childhood allergic asthma [57]. Recent studies showed that miR-206 expression was also decreased in mouse models of occupational asthma [58, 59]. In human, it was reported that circulating miR-206 was useful to predict childhood asthma exacerbation [60], and plasma miR-206 expression differed between asthmatics with higher and lower blood eosinophil counts [61]. However, the mechanism underlying the less reduction of epithelial miR-206 in type 2-high asthma relative to type 2-low asthma requires further investigation.

To explore the role of miR-206 in asthma, we verified that CD39, an ecto-nucleotidase degrading ATP, was a target of miR-206. So far, there is no studies addressing the regulation of airway CD39 in human asthma. Previous reports regarding the role of CD39 in animal asthma models were conflicting [32, 33]. We demonstrated that CD39 expression was increased in airway epithelium in human asthma and in mice sensitized and challenged with HDM. Consistent with higher expression of miR-206 in type 2-high asthma, CD39 expression was lower in type 2-high asthma relative to type 2-low asthma.

Extracellular ATP serves as a danger signal to alert the immune system of tissue damage. In our cohort of symptomatic and treatment-naïve asthma patients, BALF ATP concentrations were significantly increased, especially in type 2-high asthma patients. This is consistent with a previous report that allergen provocation enhanced airway BALF ATP accumulation in asthma patients [28]. The elevated airway ATP levels in our asthma patients might be due to exposure to environmental aeroallergen including HDM and/or airway virus infections, the main triggers of asthma exacerbation [12, 62]. It was reported that rhinovirus infection stimulated bronchial smooth muscle cells to release ATP [63], and influenza A virus infection increased BALF ATP levels in mice [64].

In our *in vitro* system, we demonstrated that HDM-induced acute extracellular ATP accumulation was responsible for miR-206 downregulation and CD39 upregulation in HBE cells on an air-liquid interface. Since CD39 catalyzes ATP degradation, the CD39 upregulation in asthma patients and in HDM-exposed HBE cells may represent an inhibitory feedback response to excessive ATP, by which striving to maintain airway homeostasis.

In human asthma, we previously reported that epithelial *IL-25* mRNA expression was upregulated in a subset of asthma patients featured by type 2 inflammation [42]. Here, in a different cohort of asthmatics, we reported that airway expression of IL-25 and TSLP whereas not IL-33 was elevated in type 2-high asthma using ELISA and qPCR. The Taqman qPCR primers and probes were previously reported for the *IL-33* isoform without exons 3 and 4, and the long isoform of *TSLP*, and these isoforms were associated with type 2 inflammation [47, 54]. So far, there have been few head-to-head studies of IL-25, IL-33 and TSLP expression in human asthma. Various expression patterns of these cytokines were reported in different populations. In Korean, plasma IL-25 whereas not IL-33 or TSLP was increased in patients with aspirin-exacerbated respiratory disease characterized by asthma, nasal polyps and chronic eosinophilic sinusitis [21]. Elevated IL-25, but not IL-33 or TSLP mRNA, was reported in sputum cells from uncontrolled asthmatics in Belgium [22]. In contrast, increased expression of IL-33 and TSLP whereas not IL-25 was reported in asthma patients in London [23], Poland [24], and New York [25]. This suggests that ethnicity and region should be considered in studies on these cytokines. Recently, a new concept “regiotype” was introduced for allergic diseases, referring to regional difference between endotypes due to different allergens and other environmental influences [2]. In chronic rhinosinusitis with nasal polyps (CRSwNP), a disease related to asthma, various expression patterns of IL-25, IL-33 and TSLP in different population and regions have been described. Reports of IL-25 upregulation in CRSwNP came from Asian countries including Korea [65], Japan [66] and China [67], whereas negative results of IL-25 were from USA [68], Australia [69] and Turkey [70]. Our data of airway IL-25, IL-33 and TSLP expression in asthma patients will provide evidence for the novel therapies targeting these cytokines in China.

The upstream signaling pathway regulating the expression of epithelial IL-25 and TSLP remains largely unknown. Here we reported that BALF ATP concentrations strongly correlated with airway IL-25 and TSLP expression. *In vitro*, allergen-induced acute accumulation of extracellular ATP was required and sufficient for IL-25 and TSLP expression. Further, epithelial miR-206 regulated IL-25 and TSLP expression by targeting CD39-extracellular ATP axis both *in vitro* and *in vivo*. HDM sensitization and challenge decreased airway miR-206 expression while increased Cd39 expression, BALF ATP concentration, Il-25, Il-33, Tslp expression, and ILC2s expansion in mice. Airway miR-206 antagonism before HDM challenge suppressed Il-25, Il-33, Tslp expression, ILC2s expansion, type 2 cytokine expression, and the cardinal features of asthma in mice. On the other hand, miR-206 overexpression had opposite effects.

As summarized in Figure 10, our findings suggest that epithelial miR-206 is downregulated in both asthma subsets. Compared with type 2-low asthma, less reduction of epithelial miR-206 resulted in higher miR-206, lower CD39 expression and impaired capacity to eliminate extracellular ATP in type 2-high asthma. Consequently, more extracellular ATP is accumulated leading to higher expression of IL-25, TSLP and more prominent type 2 inflammation in type 2-high asthma. Alternative cascades may also explain the effect of miR-206 perturbation on type 2 response in mice. For example, increased extracellular ATP can directly activate other immune cells including dendritic cells [28]. ATP can also promote dendritic cell-driven Th2 sensitization by inducing epithelial cells to release inflammatory cytokines [71, 72].

**Figure 10.**
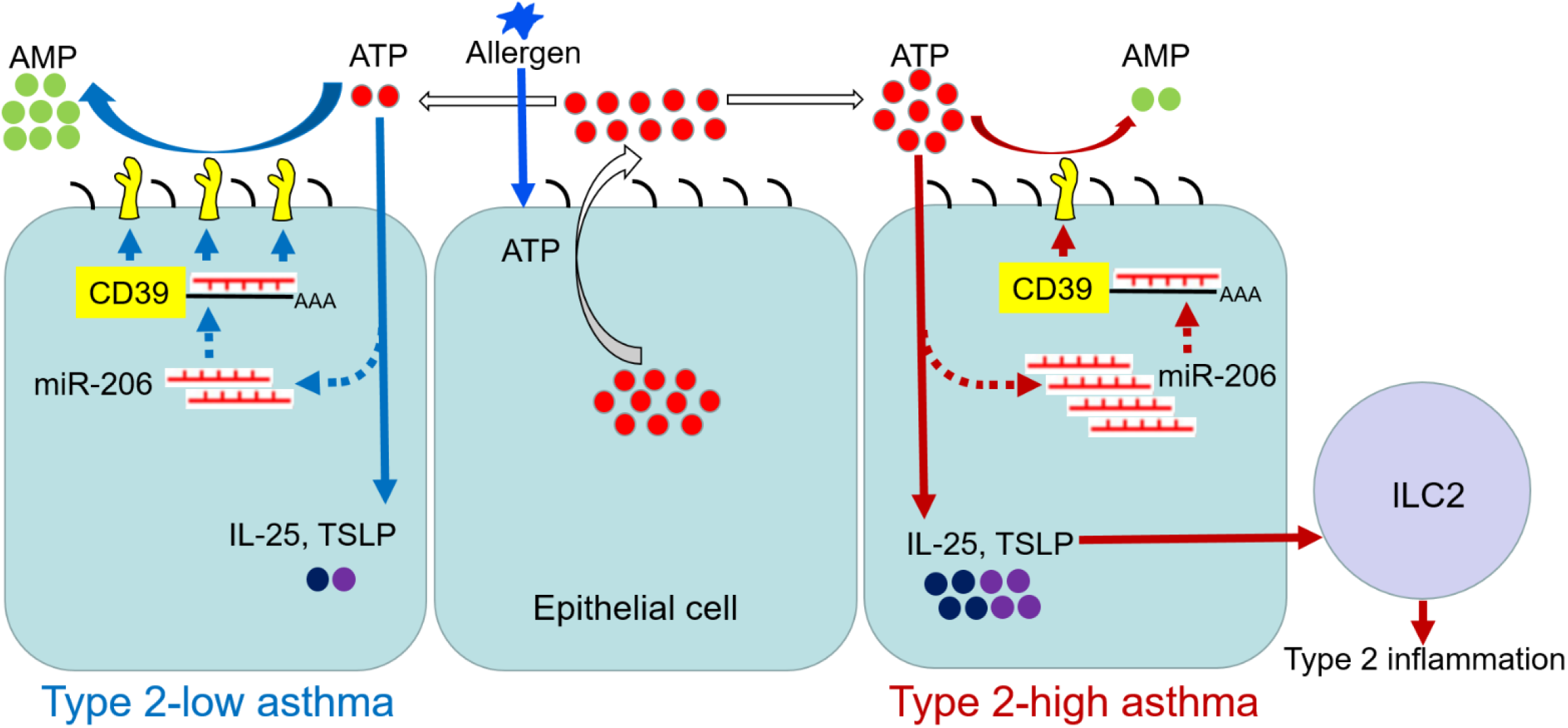
Scheme for the signaling pathway regulating IL-25 and TSLP expression in airway epithelial cells in type 2-low and type 2-high asthma. Allergen stimulates the rapid release of ATP from epithelial cells. Extracellular ATP serves as an alarmin to induce the expression of innate cytokines IL-25, TSLP. Meanwhile, acute accumulation of extracellular ATP decreases epithelial miR-206 expression, which upregulates CD39 expression to eliminate excessive ATP. Epithelial miR-206 is decreased in both type 2-low and type 2-high asthma. Compared with type 2-low asthma, less reduction of epithelial miR-206 resulted in higher miR-206, lower CD39 expression and impaired capacity to eliminate extracellular ATP in type 2-high asthma. Consequently, more extracellular ATP is accumulated which leads to higher expression of IL-25, TSLP and more prominent type 2 inflammation in type 2-high asthma.

In conclusion, epithelial miR-206 regulates airway IL-25 and TSLP expression and type 2 inflammation in asthma via targeting CD39-extracellular ATP axis. This pathway contributes, at least in part, to the development of human type 2-high asthma and represents a novel therapeutic target for this endotype.

## Data Availability

All data referred to in the manuscript are available. The array data are available at GEO (http://www.ncbi.nlm.nih.gov/geo/, accession number GSE142237).

## Acknowledgements

The authors thank all participants in this study; Xiaoling Rao and Mei Liu for bronchoscopy support; Wang Ni, Shixin Chen and Kun Zhang for spirometry measurement.

## Author contributions

G.Z. designed the research, conceived of the manuscript and had primary responsibility for writing. K.Z., Y.F., Y.L., W.W., C.C., D.C., S.C., J.G., and G.C. performed experiments. K.Z., Y.F., Y.L., L.Y., D.C., and G.Z. analyzed data. K.Z., Y.F., Y.L., L.Y., D.C., and G.Z. interpreted results of experiments. K.Z., Y.F., and G.Z. prepared figures and drafted manuscript. G.Z. edited and revised manuscript. All authors reviewed and approved the manuscript.

