## Supplementary Table and Figure for "An epithelial microRNA upregulates airway IL-25 and TSLP expression in type 2-high asthma via targeting CD39-extracellular ATP axis"

[1]Division of Pulmonary and Critical Care Medicine, Department of Internal Medicine, Tongji Hospital, Tongji Medical College, Huazhong University of Science and Technology, Wuhan, China; [2]Key Laboratory of Respiratory Diseases, National Health Commission of People's Republic of China, and National Clinical Research Center for Respiratory Diseases, Wuhan, China; [3]Department of Respiratory and Critical Care Medicine, The Affiliated Suzhou Hospital of Nanjing Medical University, Suzhou, China; [4]Department of Respiratory and Critical Care Medicine, Renmin Hospital of Wuhan University, Wuhan, China.

**Supplementary Table. Primers for quantitative PCR**

| Gene | Species | Type | Sequence |
|---|---|---|---|
| *β-actin* | Human | Forward | GCAAGCAGGACTATGACGAG |
| | | Reverse | CAAATAAAGCCATGCCAATC |
| *CD39* | Human | Forward | ACTATCGAGTCCCCAGATAATGC |
| | | Reverse | CCTGATCCTTCCCATAGCACAA |
| *CLCA1* | Human | Forward | ATGGCTATGAAGGCATTGTCG |
| | | Reverse | TGGCACATTGGGGTCGATTG |
| *GAPDH* | Human | Forward | AAGGTGAAGGTCGGAGTCAAC |
| | | Reverse | GGGGTCATTGATGGCAACAATA |
| *POSTN* | Human | Forward | GACCGTGTGCTTACACAAATTG |
| | | Reverse | AAGTGACCGTCTCTTCCAAGG |
| *SERPINB2* | Human | Forward | TCCTGGGTCAAGACTCAAACC |
| | | Reverse | CATCCTGGTATCCCCATCTACA |
| *β-actin* | Mouse | Forward | GGCTGTATTCCCCTCCATCG |
| | | Reverse | CCAGTTGGTAACAATGCCATGT |
| *Gapdh* | Mouse | Forward | TGGCCTTCCGTGTTCCTAC |
| | | Reverse | GAGTTGCTGTTGAAGTCGCA |
| *Cd39* | Mouse | Forward | AGATGAAATCGGTGCGTACCT |
| | | Reverse | GAGTCTGGTGATGCTTGGATG |

**Supplementary Figure and Figure Legend**:

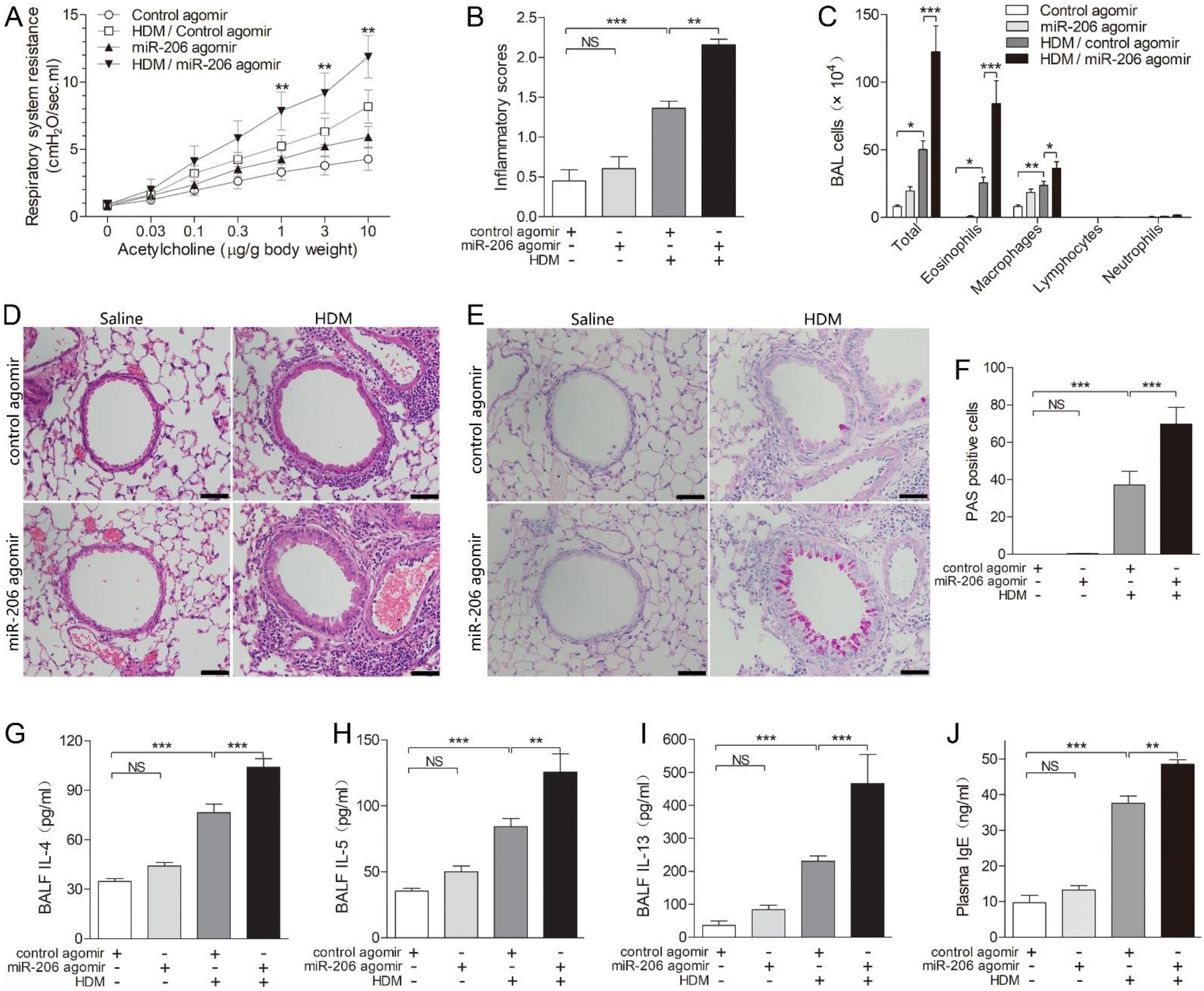


**Supplementary Figure. Overexpression of airway miR-206 expression aggravates HDM-induced AHR, airway inflammation, mucus overproduction and type 2 response in mice.** (A) Respiratory resistance in response to different concentration of intravenous acetycholine at 24 h after the last HDM or saline challenge in mice intranasally administered with control or miR-206 agomir. (B) Inflammatory scores of lung sections from mice intranasally administered with control or miR-206 agomir and challenged with HDM or saline were calculated as described in Methods. (C) Counts for macrophages, eosinophils, lymphocytes and neutrophils in BALF. (D) H&E staining of representative lung sections. (E) PAS staining for mucus in representative lung sections. (F) The numbers of PAS-staining-positive cells were counted in four random fields for each lung section at ×200 magnification. (G-I) The protein levels of IL-4 (*G*), IL-5 (*H*), IL-13 (*I*) in BALF were determined by ELISA. (J) Plasma IgE levels in peripheral blood were determined by ELISA. n = 6-10 mice per group combined from 2 experiments. Data are mean ± SD. *$P$<0.05; **$P$<0.01; ***$P$<0.001 (one-way ANOVA with Bonferroni's post hoc test). Scale bar = 50 μm.
